# Investigating Causal Associations of Diet-Derived Circulating Antioxidants with Risk of Six Major Mental Disorders: A Mendelian Randomization Study

**DOI:** 10.1101/2022.05.11.22274935

**Authors:** Hao Zhao, Xue Han, Lingjiang Li, Xuening Zhang, Yuhua Liao, Huimin Zhang, Wenyan Li, Jingman Shi, Wenjian Lai, Wanxin Wang, Roger S. McIntyre, Kayla M. Teopiz, Lan Guo, Ciyong Lu

**Affiliations:** Department of Medical Statistics and Epidemiology, School of Public Health, Sun Yat-sen University, Guangzhou, China; Guangdong Provincial Key Laboratory of Food, Nutrition and Health, Sun Yat-sen University, Guangzhou, China; Department of Psychiatry, Shenzhen Nanshan Center for Chronic Disease Control, ShenZhen, China; Mental Health Institute of the Second Xiangya Hospital, Central South University, Changsha, China; Department of Epidemiology and Biostatistics, College of Public Health, Shandong University, Jinan, China; Department of Pharmacology, University of Toronto, Toronto, Ontario, Canada; Department of Psychiatry, University of Toronto, Toronto, Ontario, Canada; Mood Disorders Psychopharmacology Unit, University Health Network, Toronto, Ontario, Canada; Institute of Medical Science, University of Toronto, Toronto, ON, Canada

**Keywords:** Antioxidant, α-Tocopherol, Schizophrenia, Mental disorder, Mendelian randomization, Depression, Bipolar disorder, Autism spectrum disorder, Attention-deficit/hyperactivity disorder, Post-traumatic stress disorder, Non-communicable diseases, Public health

## Abstract

**Background:** Observational studies have suggested associations between circulating antioxidant levels and many mental disorders, but evidence from randomized controlled trials (RCTs) is lacking and causal inferences have not been confirmed. The aim of this study was to explore whether genetically predicted diet-derived circulating antioxidants were causally associated with the risk of major mental disorders using Mendelian randomization (MR).

**Methods and findings:** We performed 2-sample MR analyses of summary-level genetic data to explore whether diet-derived circulating antioxidants [e.g., vitamins E (α- and γ-tocopherol), ascorbate, retinol, β-carotene, and lycopene], assessed by absolute circulating antioxidants and relative circulating antioxidant metabolites, were causally associated with the risk of six major mental disorders, including major depressive disorder (MDD), schizophrenia (SCZ), bipolar disorder (BIP), autism spectrum disorder (ASD), attention-deficit/hyperactivity disorder (ADHD), and post-traumatic stress disorder (PTSD). The inverse-variance weighted method was adopted as primary MR analyses and five additional MR methods (likelihood-based MR, MR-Egger, weighted median, penalized weighted median, and MR-PRESSO) and different outcome databases were used for sensitivity analyses. We found suggestive evidence that genetically predicted higher absolute circulating α-tocopherol levels marginally reduced the risk of SCZ, with the odds ratio (OR) per unit increase in log-transformed α-tocopherol values was 0.71 [95% confidence interval (CI) 0.54 to 0.94; *P* = 0.016]. However, after adjusting for multiple testing (threshold of *P* < 0.008), we found no significant evidence that genetically predicted higher diet-derived absolute circulating antioxidant levels and antioxidant metabolites concentrations were significantly causally associated with the six-foregoing major mental disorders.

**Conclusions:** Overall, our study does not support significant causal associations of genetically predicted diet-derived circulating antioxidants with the risk of major mental disorders. Therefore, simply taking antioxidants to increase blood antioxidants levels is unlikely to have a significant protective effect on the prevention of most mental disorders.

**Author summary:** *Why was this study done?:* Some observational studies have reported that diet-derived circulating antioxidants are associated with a reduced risk of major mental disorders; however, these studies are susceptible to uncertain temporal relationships, insufficient sample sizes, or potential confounding factors, and thus it remains unclear whether these associations are accurate. To our knowledge, there are no randomized clinical trials published to date on this topic. Since oxidative stress is closely related to the occurrence of mental diseases, if diet-derived circulating antioxidants can reduce the risk of major mental disorders, it will be an interesting target as primary prevention of mental disorders.

*What did the researchers do and find?:* We performed a Mendelian randomization study design to explore whether genetically predicted diet-derived circulating antioxidants [e.g., vitamins E (α- and γ-tocopherol), ascorbate, retinol, β-carotene, and lycopene], assessed by absolute circulating antioxidants and relative circulating antioxidant metabolites, were causally associated with the risk of six major mental disorders, including major depressive disorder, schizophrenia, bipolar disorder, autism spectrum disorder, attention- deficit/hyperactivity disorder, and post-traumatic stress disorder. Overall, our study provides suggestive evidence that genetically predicted higher absolute α-tocopherol levels may be causally associated with a reduced risk of schizophrenia. However, our study did not find genetically predicted significant causal associations of dietary antioxidants with major mental disorders after correction for multiple testing.

*What do these findings mean?:* Our findings suggest for healthy adults without nutritional deficiency, simply taking antioxidants to increase blood antioxidants levels is unlikely to have a significant protective effect on the prevention of most mental disorders. In the future, large-scale GWASs are needed to further validate our current findings, especially the suggestive protective effect of higher α-tocopherol levels on schizophrenia, by utilizing additional genetic variants and more samples.

## Introduction

With the acceleration of the pace of life, the intensification of competitive pressure, and the influence of COVID-19, people’s psychological pressure is increasing, and the prevalence of mental disorders is increasing year by year [1]. Major mental disorders such as major depressive disorder (MDD), schizophrenia (SCZ), bipolar disorder (BIP), autism spectrum disorder (ASD), attention- deficit/hyperactivity disorder (ADHD), and post-traumatic stress disorder (PTSD), constitute a large part of global health burden worldwide [2]. Mental disorders are caused by a variety of complex factors, including genetic, stress, biological, psychological, and environmental factors. Although considerable efforts have been made to understand the nature of mental disorders, understanding of their pathogenesis remains limited, and there are no effective etiological prevention methods. It has been reported that oxidative stress can cause oxidative damage to biological macromolecules, cells, and neurons, which is considered to be one of the main pathogenesis of mental disorders [3]. Moreover, it has been reported that antioxidants, which can help eliminate free radicals and reduce and eliminate oxidative damage [4], would be an interesting target as primary prevention of mental disorders. It is worth noting that antioxidants from dietary intake are the most easily accessible and modifiable approach for consideration.

Research on the relationship between antioxidants and major mental disorders has never stopped. Some observational studies have shown that dietary intake, either as dietary components or supplements, or the concentration of vitamins E and C, and carotenoids in the blood, are associated with a reduced risk of MDD [5-7], SCZ [8, 9], BIP [10], ASD [11, 12], ADHD [13], and PTSD [14]. However, some observational studies do not report a protective effect of the foregoing antioxidants on mental disorders [15-19]. The inconsistent results reported in some observational studies may be due to uncertain temporal relationships, insufficient sample sizes, short follow-up periods, or potential confounding factors. Previous clinical trials of antioxidants have focused more on improving symptoms in people with mental disorders [20, 21], and there is still a lack of randomized clinical trials (RCTs) investigating whether diet-derived antioxidants can prevent mental disorders. However, intervention studies in healthy people cannot be conducted without sufficient evidence because of the potential for unknown risks and harms to the subjects. Moreover, intervention trials are often limited by timing, dosage, duration, use of natural or synthetic antioxidants, and the uncertainty of the onset time and long-term progression of mental disorders. Therefore, the causal relationship between diet-derived antioxidants and the risk of major mental disorders remains unclear.

Mendelian randomization (MR), which uses genetic variants of the exposure as instrumental variables to minimize measurement errors, confounding, and reversed causation, can provide reliable estimation of the causal association between exposure and outcome under specific assumptions [22]. With the development of sequencing technology, many large-scale genome-wide association studies (GWAS) data on diet-derived antioxidants and major mental disorders have been published, which provides an opportunity for 2-sample MR analyses using summary statistics from separate studies to substantially increase the statistical power by combining data from multiple sources.

In this study, we used a 2-sample MR analysis to assess the causal associations between genetically predicted diet-derived circulating antioxidants and major mental disorders, including MDD, SCZ, BIP, ASD, ADHD, and PTSD.

## Methods

### Overall study design

The study herein used 2-sample MR analyses of summary level genetic data to investigate whether diet-derived circulating antioxidants, including vitamins E (α- and γ-tocopherol), ascorbate, retinol, β-carotene, and lycopene, were causally associated with the risk of six major mental disorders, including MDD, SCZ, BIP, ASD, ADHD, PTSD. We considered the following two phenotypes for these antioxidants: (1) absolute circulating antioxidants: authentic absolute levels measured by targeting in the blood, (2) circulating antioxidant metabolites: relative concentrations quantified by untargeted metabolomics in plasma or serum, either or both. The instrumental variables need to satisfy three assumptions: the relevance assumption, the independence assumption, and the exclusion restriction assumption [23]. This study is reported according to the Strengthening the Reporting of Observational Studies in Epidemiology Using Mendelian Randomization (STROBE-MR) checklist (S1 STROBE-MR-Checklist).

### Determination of exposures

For genetic instrumental variables of absolute circulating antioxidants, genetically predicted α- tocopherol, ascorbate, retinol, β-carotene, and lycopene were identified in the recent large-scale GWAS analysis (*P* < 5e-8, linkage disequilibrium [LD]: r^2^ < 0.001 and clump distance = 10,000 kb). Three independent single nucleotide polymorphisms (SNPs) associated with α-tocopherol were identified from a GWAS with 4,014 individuals of European ancestry [24]. Ten independent SNPs associated with ascorbate were identified from a recently published GWAS with 52,018 individuals of European ancestry [25]. Two independent SNPs associated with retinol were identified from a GWAS of 5,006 Caucasian individuals in two cohort studies [26]. Two independent SNPs associated with β- carotene were identified from a GWAS of 2,344 participants in the Nurses’ Health Study [27]. Five independent SNPs associated with lycopene were identified from a GWAS with 441 Caucasian participants [28].

For genetic instrumental variables of circulating antioxidant metabolites, genetically predicted α- tocopherol, γ-tocopherol, ascorbate, and retinol were extracted from the metabolite GWAS analysis at suggestive genome-wide significance level (*P* < 1e-5). A total of 11 SNPs for α-tocopherol (n = 7,725), 13 SNPs for γ-tocopherol (n = 6,226), and 14 SNPs for ascorbate (n = 2,085) were derived from 7,824 adult individuals from two European population studies [29], and 26 SNPs for retinol from 1,960 adults of European descent [30].

Based on the PhennoScanner database, the SNPs not reaching the *P* < e-5 level of association with the confounders remained in the filtered genetic instrument [31]. The proportion of variance explained (R^2^) of instrument for each exposure was either derived from the original study or calculated based on the derived summary statistics by the following formula: R^2^ = (2 × EAF × (1 - EAF) × Beta^2^) / [(2 × EAF × (1 - EAF) × Beta^2^) + (2 × EAF × (1 - EAF) × N × SE^2^)]. Beta indicates the estimated genetic effect of SNP on each exposure, EAF is effect allele frequency, SE is standard error of the estimated effect and N is sample size. Then, the F statistic for each SNPs was calculated by the following formula: Beta^2^ / SE^2^, and the F-statistics of >10 for each SNP is recommended to avoid employing week genetic instruments.

### Data sources of major mental disorders

The latest GWAS summary statistics for MDD, SCZ, BIP, ASD, ADHD, and PTSD were extracted from the Psychiatric Genomics Consortium (PGC) website (https://www.med.unc.edu/pgc/results-and-downloads/). The PGC is the largest consortium in the history of psychiatry, which has conducted the most influential meta- and mega-analysis of genome-wide genomic data for mental disorders. The samples were collected from multiple cohorts, mostly of European ancestry. The GWAS summary datasets for MDD (170,756 cases and 329,443 controls) in 2019 [32], SCZ (67,390 cases and 94,015 controls) in 2021 [33], BIP (41,917 cases and 371,549 controls) in 2021 [34], ASD (18,382 cases and 27,969 controls) in 2019 [35], ADHD (20,183 cases and 35,191 controls) in 2019 [36], and PTSD (23,212 cases and 151,447 controls) in 2019 [37] were obtained from genome-wide meta- or mega-analysis. For SNPs of instrument were not available in the outcome GWAS, the LDlink tool was used to identify proxy SNPs of European ancestry (r^2^ > 0.8) [38]. SNPs missing in the outcome GWAS without appropriate proxy SNPs available were then excluded.

### Statistical analysis

The selection of main MR analyses methods is shown in ***Figure 1***. First, the MR-Egger intercept test was performed to test whether there was the presence of potential pleiotropy [39]. If there was significant horizontal pleiotropy, the MR-Egger regression was used, otherwise the inverse-variance weighted (IVW) meta-analysis was used, which assumes that either all the instruments are valid or any horizontal pleiotropy is balanced. Then, heterogeneity was assessed using Cochran’s Q-statistics test [40]. If there was significant heterogeneity, the random-effect IVW model was used, otherwise the fixed-effect IVW model was used.

**Figure 1.**
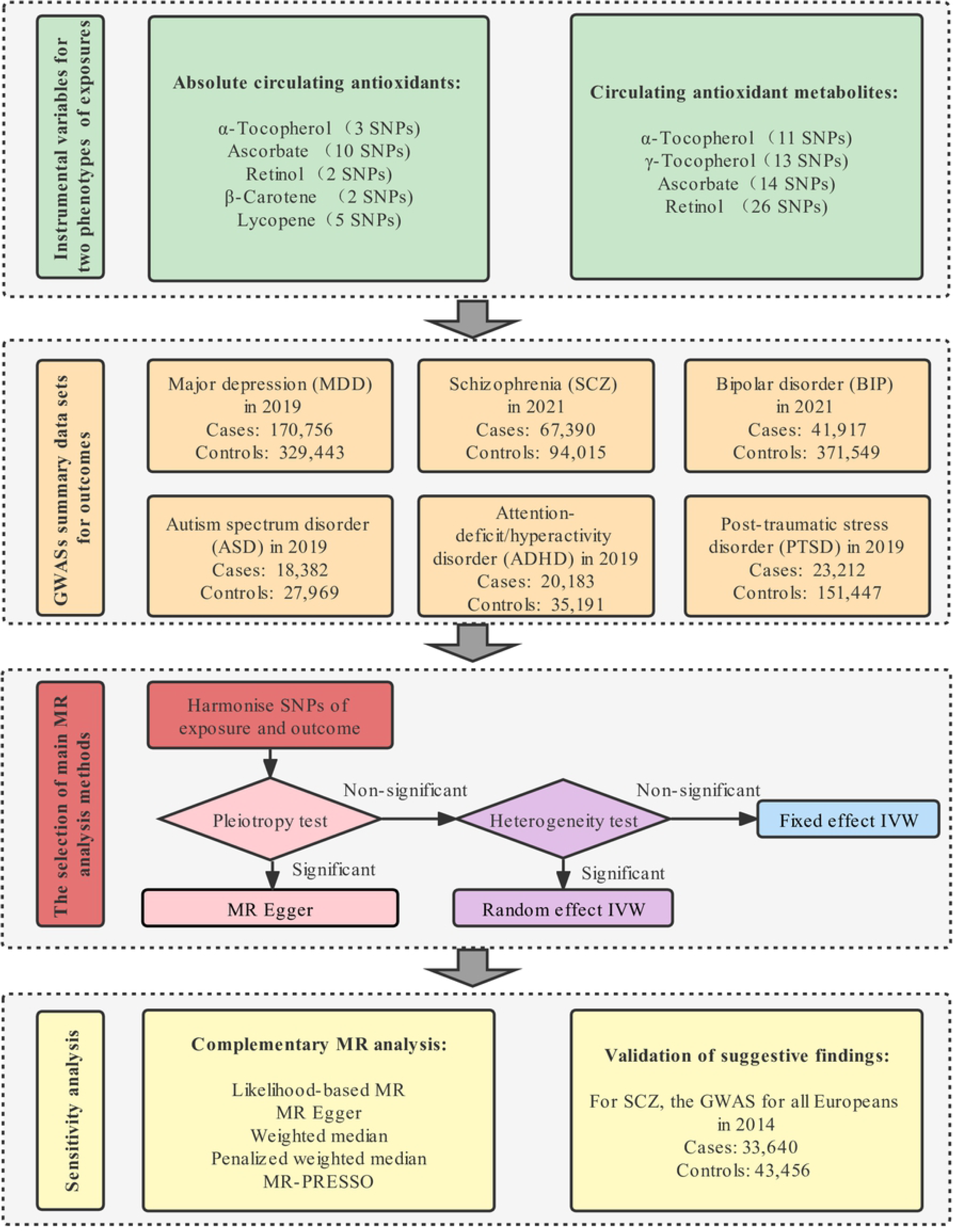
Study design and framework of this research. The pleiotropy test refers to the MR-Egger intercept test, and the heterogeneity test refers to the Cochran’s Q-statistics test.

To further assess the robustness of our findings, a series of sensitivity analyses were performed. First, complementary MR analyses with different assumptions were applied to help verify causal inference. The likelihood-based MR was considered the most accurate method to estimate causal effects when there was a continuous log-linear association between exposure and the risk of outcome (No. SNPs > 1) [41]. The weighted median can provide valid estimates if at least 50% of the weight comes from valid instrumental variables (No. SNPs > 2) [42]. The penalized weighted median method was implemented, which derives valid causal estimates even under conditions when invalid instruments are present (No. SNPs > 2) [42]. The MR Pleiotropy RESidual Sum and Outlier (MR- PRESSO) was performed, which detects and corrects the effects from outliers (No. SNPs > 3) [43]. Second, we also performed scatter, forest, and leave-one-out plots to detect high influence points. Third, the discovery of suggestive evidence was further verified in 2014 GWAS results of SCZ in the European population (33,640 cases and 43,456 controls) [44].

The priori statistical power was calculated using the mRnd power calculation online tool [45]. Given a type 1 error of 5%, we had sufficient power (> 80%) to detect the minimum detectable odds ratio (OR) per unit increase of antioxidants. To account for multiple testing in our analyses, we used a Bonferroni-corrected threshold of *P* < 0.008 (α = 0.05/6 outcomes) as significant evidence of associations, and a *P*-value between 0.05 and 0.008 was considered suggestive evidence of associations. All statistical analysis were performed using R version 4.1.0.

## Results

### Strength of genetic instruments

The summary information of GWAS for diet-derived absolute circulating antioxidants and metabolites and major mental disorders is shown in ***Table 1*** and ***S1 and S2 Tables***. The genetic instruments of α- tocopherol, ascorbate, and retinol were available both as absolute circulating antioxidants and metabolites. Variance explained by the genetic instruments ranged from 1.7% to 30.1% for absolute circulating antioxidants (all F statistic >10) and from 6.8% to 21.7% for antioxidants metabolites (all F statistic >10). The statistical power in MR study suggested that for most analyses we had the adequate statistical power to identify even modest causality (***S3 Table***). The raw data information on the effect estimation for the associations of selected SNPs with antioxidants and with major mental disorders is given in ***S4 and S5 Tables***.

**Table 1.** The summary of instrumental variables for diet-derived absolute circulating antioxidants and antioxidant metabolites.

| Trait | Sample size | P-value | LD | No. of SNPs | Explained variance * | Unit | PMID |
| --- | --- | --- | --- | --- | --- | --- | --- |
| **Absolute circulating antioxidants** |  |  |  |  |  |  |  |
| α-Tocopherol | 4,014 | 5e-8 | 0.001 | 3 | 1.7 | mg/L in log-transformed scale | 21729881 |
| Ascorbate | 52,018 | 5e-8 | 0.001 | 10 | 1.7 | μmol/L | 33203707 |
| Retinol | 5,006 | 5e-8 | 0.001 | 2 | 2.3 | μg/L in ln-transformed scale | 21878437 |
| β-Carotene | 2,344 | 5e-8 | 0.001 | 2 | 4.8 | μg/L in ln-transformed scale | 23134893 |
| Lycopene | 441 | 5e-8 | 0.001 | 5 | 30.1 | μg/dL | 26861389 |
| **Circulating antioxidant metabolites** |  |  |  |  |  |  |  |
| α-Tocopherol | 7,725 | 1e-5 | 0.001 | 11 | 6.8 | log10-transformed metabolites concentration | 24816252 |
| γ-Tocopherol | 6,226 | 1e-5 | 0.001 | 13 | 9.8 | log10-transformed metabolites concentration | 24816252 |
| Ascorbate | 2,085 | 1e-5 | 0.001 | 14 | 21.7 | log10-transformed metabolites concentration | 24816252 |
| Retinol | 1,960 | 1e-5 | 0.001 | 26 | 20.6 | log10-transformed metabolites concentration | 28263315 |

LD: linkage disequilibrium.
\* Variance explained ( $R^2$ ) were either derived from the original study or calculated $R^2 = (2 \times \text{EAF} \times (1 - \text{EAF}) \times \text{Beta}^2) / [(2 \times \text{EAF} \times (1 - \text{EAF}) \times \text{Beta}^2) + (2 \times \text{EAF} \times (1 - \text{EAF}) \times N \times \text{SE}^2)]$ , where Beta indicates the estimated genetic effect of SNP on each exposure, EAF is effect allele frequency, SE is standard error of the estimated effect and N is sample size.

### Effect of absolute circulating antioxidants on the risk of major mental disorders

Primary results of MR estimate for absolute circulating antioxidants are presented in ***Figure 2***. For absolute circulating antioxidants with 3 or more SNPs, the MR-Egger intercept test found no significant horizontal pleiotropy for all outcomes, with *P*-values ranging from 0.09 to 0.83. According to the IVW method, we found suggestive evidence that genetically predicted higher absolute α- tocopherol levels reduced the risk of SCZ, with the OR per unit in log-transformed α-tocopherol values was 0.71 (95% confidence interval [CI] 0.54 - 0.94; *P* = 0.016). However, after the Bonferroni-corrected significance threshold, we observed no significant evidence that genetically predicted absolute circulating antioxidant levels were significantly associated with the six major mental disorders (all *P* > 0.008).

**Figure 2.**
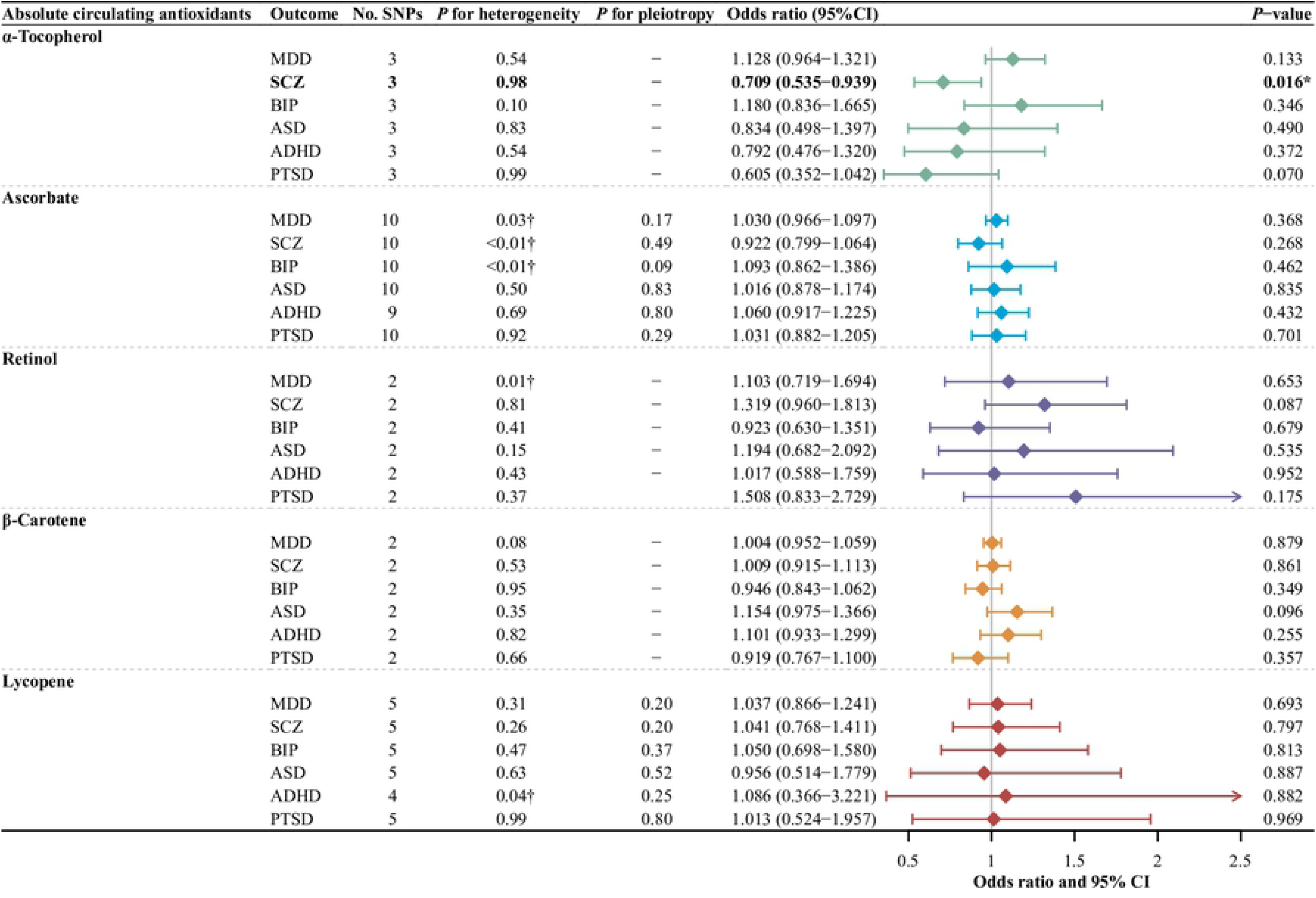
The main MR analysis results of the causal effects of absolute circulating antioxidant levels on six major mental disorders. (†) means there is significant heterogeneity (*P* for heterogeneity < 0.05), using the random-effect IVW model, otherwise using the fixed-effect IVW model. The odds ratios are scaled per unit increase in log-transformed α-tocopherol values, per unit increase in ascorbate, per unit increase in ln-transformed retinol and β-carotene values, and per 10 unit increase in lycopene. Statistical significance was defined as Bonferroni-corrected threshold of *P* < 0.008 (0.05/6), and *P*-value between 0.05 and 0.008 was considered suggestive evidence (*) of associations. MDD: major depressive disorder, SCZ: schizophrenia, BIP: bipolar disorder, ASD: autism spectrum disorder, ADHD: attention-deficit/hyperactivity disorder, PTSD: post-traumatic stress disorder.

### Effect of circulating antioxidant metabolites on the risk of major mental disorders

***Figure 3*** shows the primary results of MR estimates for circulating antioxidant metabolites. Similarly, the MR-Egger intercept test also found no significant horizontal pleiotropy for all outcomes, with *P*- values ranging from 0.11 to 0.97. Similar with the findings from analyses of absolute circulating antioxidants, after the Bonferroni-corrected significance threshold, we also observed no significant evidence that genetically predicted circulating antioxidant metabolites were significantly associated with the six major mental disorders (all *P* > 0.008).

**Figure 3.**
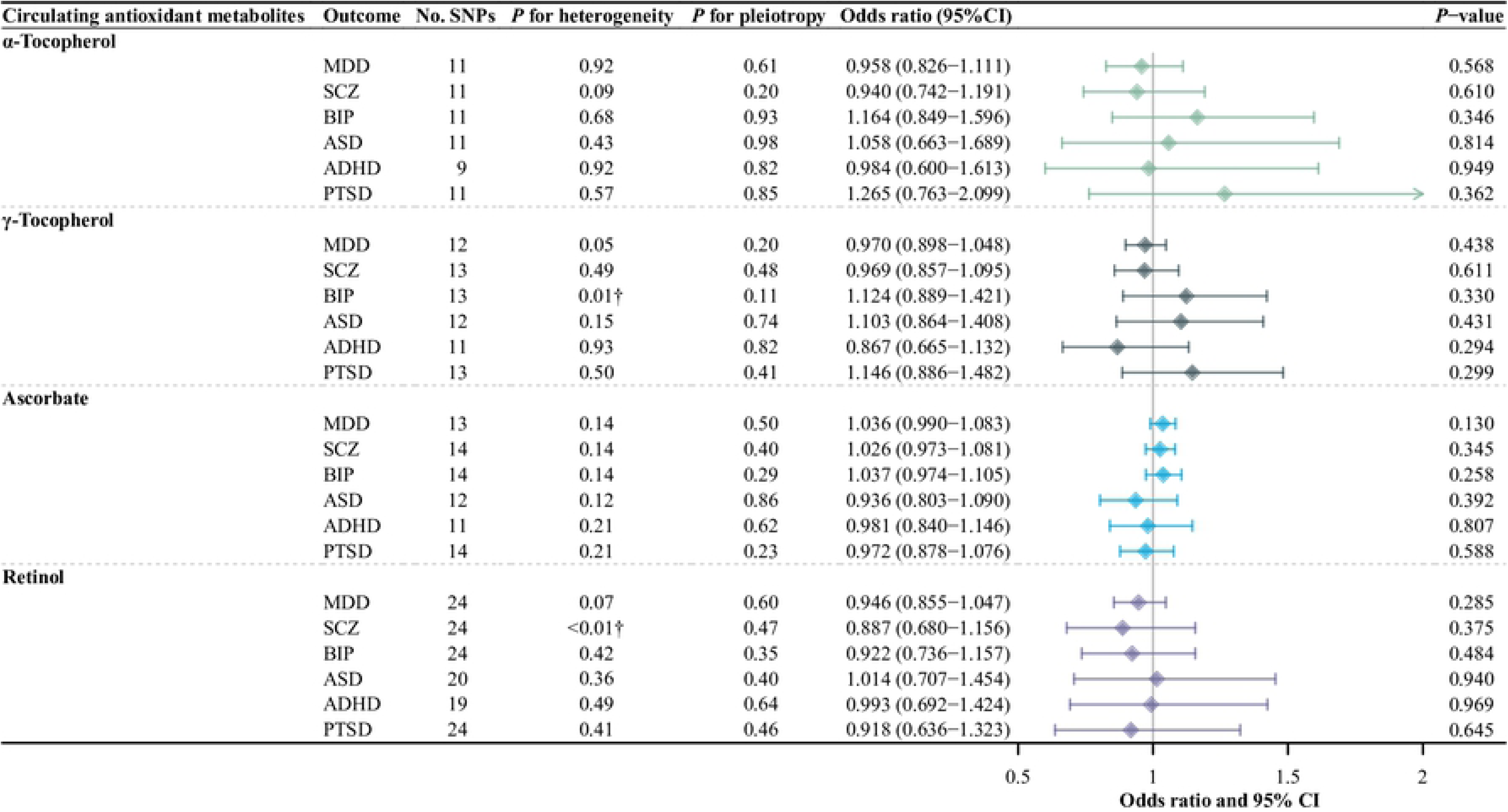
The main MR analysis results of the causal effects of circulating antioxidant metabolites on six major mental disorders. (†) means there is significant heterogeneity (*P* for heterogeneity < 0.05), using the random-effect IVW model, otherwise using the fixed-effect IVW model. The odds ratios are scaled per unit increase in log-transformed α-tocopherol, γ-tocopherol, and ascorbate values, and per 10 unit increase in log- transformed retinol values. Statistical significance was defined as Bonferroni-corrected threshold of *P* < 0.008 (0.05/6), and *P*-value between 0.05 and 0.008 was considered suggestive evidence (*) of associations. MDD: major depressive disorder, SCZ: schizophrenia, BIP: bipolar disorder, ASD: autism spectrum disorder, ADHD: attention- deficit/hyperactivity disorder, PTSD: post-traumatic stress disorder.

### Sensitivity analysis

For the suggestive evidence of the protective effect of absolute α-tocopherol levels on SCZ in the primary analysis (IVW method), the supplementary MR analyses showed that, genetically determined higher absolute α-tocopherol levels marginally were associated with the lower risk of SCZ based on the likelihood-based MR (OR 0.71, 95% CI 0.53 - 0.95, *P* = 0.023, ***Figure 4***). The scatter, forest, and leave-one-out plots showed that although rs2108622 may weaken the effect (because there were only three SNPs for instrumental variables of absolute α-tocopherol levels), no single SNP having a large effect on the overall results (***S 1-3 Figure***). The leave-one-out test also suggested that the results were robust even though rs964184 might have been eliminated due to its association with cholesterol and triglycerides. In addition, we further confirmed that higher absolute diet-derived α-tocopherol levels marginally reduced risk of SCZ by outcome from another GWAS from all-European population (OR 0.67, 95% CI 0.45 - 1.00, *P* = 0.051, nearly reaching suggestive evidence, ***S6 Table***).

**Figure 4.**
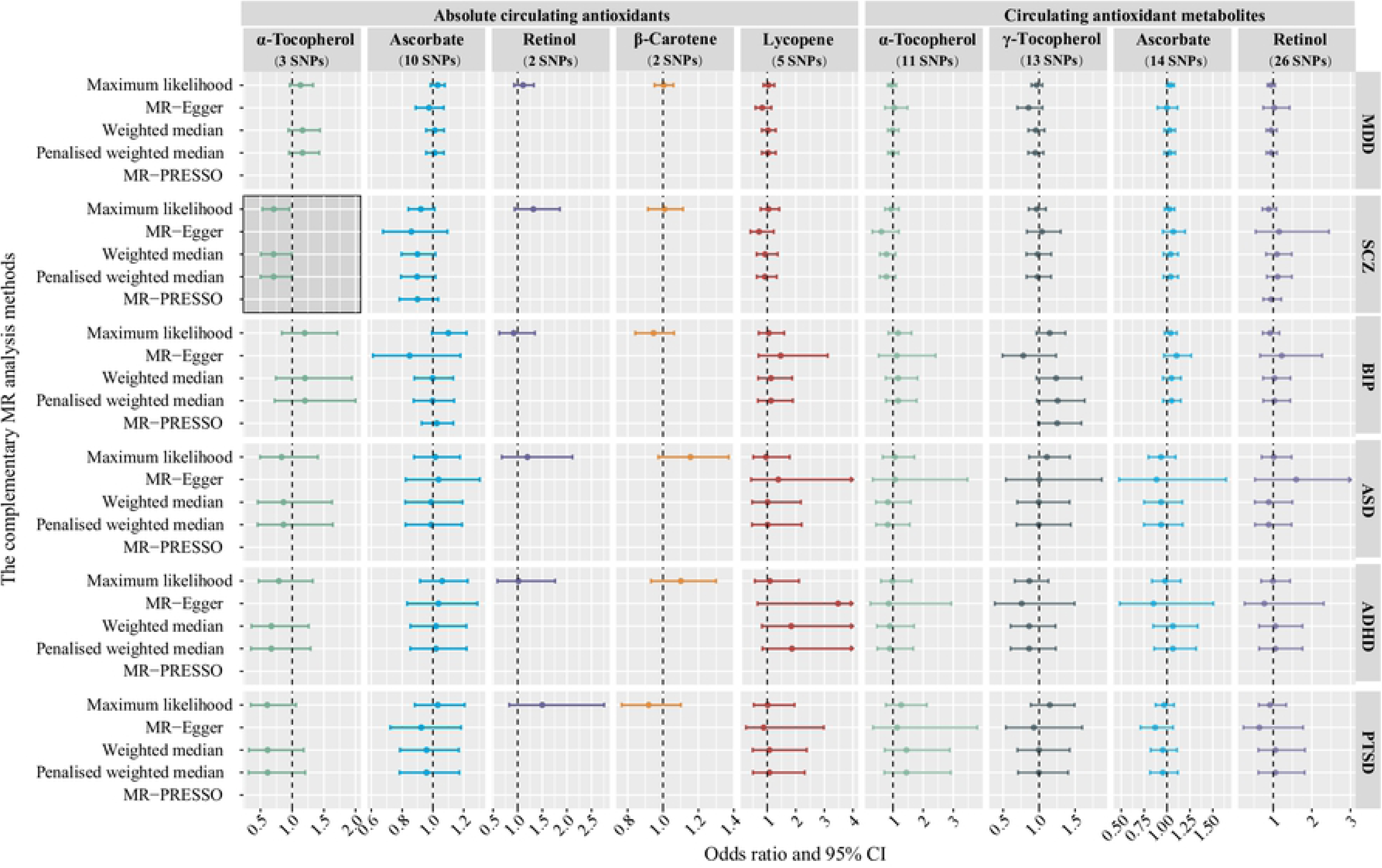
The complementary MR analyses results of the causal effects of diet-derived circulating antioxidants on six major mental disorders. The MR-Egger, weighted median and penalized weighted median require the number of SNPs in the instrumental variable > 2, and MR-Egger method cannot accurately estimate due to collinearity in MR analyses for α-tocopherols. The MR-PRESSO requires the number of SNPs in the instrument variable > 3. The MR-PRESSO was not performed because the MR-PRESSO global test did not identify significant outliers in the genetic instrument and therefore did not require correction. Thus, the causal estimate calculated by MR-PRESSO was the same as that by the IVW method. The error bars indicate 95% CIs. Shaded areas represent suggestive evidence. MDD: major depressive disorder, SCZ: schizophrenia, BIP: bipolar disorder, ASD: autism spectrum disorder, ADHD: attention-deficit/hyperactivity disorder, PTSD: post-traumatic stress disorder.

For robust null results in the primary analysis (IVW method), similar OR estimates as in the primary MR analysis but with lower precision were obtained from MR-Egger, weighted median, and penalized weighted median analysis. The MR-PRESSO method identified outlier SNP for absolute ascorbate on SCZ and BIP, for α-tocopherol metabolite on BIP, and for retinol metabolite on SCZ. Outlier-correction did not materially change the OR estimates. No outlier SNPs were identified in the MR-PRESSO analysis for the other outcomes (***Figure 4***).

## Discussion

In this study, we investigated the causal associations between diet-derived circulating antioxidants and the risk of six major mental disorders based on MR analyses. The genetic variation of circulating antioxidants was evaluated as authentic absolute blood levels and relative metabolites concentrations as instrumental variables, and comparable results were obtained. We only found suggestive evidence that genetically predicted higher absolute circulating α-tocopherol levels marginally reduced the risk of SCZ. However, we found no significant evidence that genetically predicted higher diet-derived antioxidants were significantly causally associated with the six major mental disorders after correction for multiple testing.

An case-control study by Dadheech et al. in persons at the age of 18-60 years found that the level of α-tocopherol in the blood of SCZ patients was significantly lower than that in the control group (case: 0.54 mg / dl vs control: 0.82 mg / dl, *P* < 0.001) [8]. A five-year follow-up study of fifty-five SCZ patients also observed that SCZ patients with stable phase had lower α-tocopherol levels than healthy control [46]. A study by Liu et al. which aimed to identify novel biomarkers of SCZ in peripheral blood mononuclear cells reported that SCZ subjects had significantly lower mean levels of α-tocopherol and highlighted the value of α-tocopherol in the diagnosis of SCZ [47]. The foregoing studies are consistent with our research. Our research also suggested that higher absolute α-tocopherol levels marginally reduced the risk of SCZ. Although we found that the direction of association with SCZ risk was consistent in α-tocopherol and γ-tocopherol metabolites, these associations did not reach suggestive evidence thresholds, which may be due to the lack of standards for untargeted metabolomics, resulting in many false positive signals, leading to biased results [48].

Previous studies have demonstrated that the effect of genetic variants on circulating antioxidant levels are generally comparable with those which would be achieved by dietary supplementation [49]. However, to date, although an RCT study on the prognosis of patients with SCZ has shown that vitamin E supplementation is beneficial to the treatment of SCZ [50], there is still a lack of RCT study on whether vitamin E supplementation can prevent SCZ. It is noteworthy that the effects of genetic susceptibility are lifelong, whereas the effects of antioxidant supplementation may only last up to the trial period. Considering that short-term supplementation therapy may not alter long-term risk, slight exposure throughout life would have greater potential biological effects than the temporary and large dose supplement.

The mechanisms of the suggestively potential beneficial effect of α-tocopherol on SCZ are complex. There is increasing evidence that the pathophysiology of patients with SCZ may involve excessive free radical production or oxidative stress, manifested as increased production of reactive oxygen species or decreased antioxidant protection [51-53]. Studies have shown that among various vitamin E components, only α-tocopherol is actively absorbed by the brain and directly involved in the protection of nerve membranes [54]. α-Tocopherol with lipotropy hydrophobic structure, able to enter the biofilm containing unsaturated fatty acids, can directly capture free radicals in the lipid medium and effectively terminate the chain reaction of free radicals leading to lipid peroxidation, so as to maintain the stability of the composition of unsaturated fatty acids on the membrane [55].

However, there was no significant causal relationship between genetically predicted higher diet-derived antioxidants with the major mental disorders. This seems to violate the oxidative stress hypothesis, but circulating antioxidant levels may not represent antioxidant capacity, and that increasing the level of an antioxidant in blood through simple nutritional intake or supplements does not necessarily produce additional antioxidative effects [56]. Therefore, for healthy adults without nutritional deficiency, dietary supplements that are to increase antioxidant concentrations in blood is unlikely to have a significant protective effect on the prevention of most mental disorders.

There are three main strengths in the present study. First, the MR design of two independent samples based on genetic instrumental variables reduces the possibility of subjects exposed to unnecessary risks and hazards in RCT study, and supplements the genetic theoretical basis of dietary antioxidants and major mental disorders. Second, we used genetic instrument variables based on two phenotypes, including absolute blood levels of antioxidants measured quantitatively by targeted and relative metabolite concentrations detected by untargeted metabolomics. Untargeted metabolomics is primarily used for broad coverage of metabolites, but its measurement accuracy is lower than targeted quantitative measurements due to increased false positive rates. Importantly, although there is some measurement bias for relative antioxidants metabolite concentrations, the direction of effect for the suggestive findings at the absolute levels is consistent, which also adds to the reliability of our results. Third, we further validated the robustness of suggestive findings through complementary MR analyses methods and outcome from different GWAS sources.

However, some limitations should also be considered with respect to the interpretation of the results. First, our analyses are based on published summary data rather than individual data, therefore, we were unable to test nonlinear causal relationships between antioxidant levels and risk for select mental disorders. Second, although an inherent limitation of MR analyses is that there may be potential polymorphic effects, the MR-Egger intercept test in our study has shown no significant pleiotropy, and no SNPs significantly associated with confounding were detected in the PhennoScanner database [31], indicating that horizontal pleiotropy is unlikely to exist. Third, although we only detected the weak protective effects of α-tocopherol on SCZ, and did not detect the causal association with major mental disorders for other dietary sources of antioxidants, the possibility that the effect size was too small to be recognized cannot be completely ruled out. For example, higher circulating lycopene, β-carotene and vitamin C concentrations may still be beneficial to specific populations with risk factors at baseline or oxidative stress. Fourth, our instrumental variable for α-tocopherol only contains 3 SNPs, explaining 1.7% of the variance. Although the statistical power of the MR analysis has been met, a larger GWAS is still needed to find more SNPs that may be significantly associated with α-tocopherol for further verification. Finally, due to the availability of data, this study focused on populations of European ancestry and other populations that need further validation. Future studies assessing the relationship between dietary antioxidants and risk for select mental disorders among other populations are needed for need additional supportive evidence and validation of our findings reported herein.

## Conclusions

Overall, our study provides suggestive evidence that genetically predicted higher absolute α- tocopherol levels may be causally associated with a reduced risk of SCZ. However, our study did not find genetically predicted significant causal associations of dietary antioxidants with major mental disorders after correction for multiple testing. Therefore, for healthy adults without nutritional deficiency, simply taking antioxidants to increase blood antioxidants levels is unlikely to have a significant protective effect on the prevention of most mental disorders. However, the findings do not exclude the possibility that there may be smaller effects than we could not detect. In the future, large-scale GWASs are needed to further validate our current findings by utilizing additional genetic variants and more samples.

## Data Availability

All relevant data are within the manuscript and its Supporting Information files. The summary-level data for diet-derived circulating antioxidants are available at the corresponding original study (see References 23-29). The summary-level data for six major mental disorders are available at the Psychiatric Genomics Consortium (PGC) website (https://www.med.unc.edu/pgc/results-and-downloads/).

## Data Availability Statement

All relevant data are within the manuscript and its Supporting Information files. The summary-level data for diet-derived circulating antioxidants are available at the corresponding original study [24-30]. The summary-level data for six major mental disorders are available at the Psychiatric Genomics Consortium (PGC) website (https://www.med.unc.edu/pgc/results-and-downloads/).

## URLs

PGC, http://www.med.unc.edu/pgc/;

GWAS Catalog, https://www.ebi.ac.uk/gwas/;

IEU OpenGWAS project, https://gwas.mrcieu.ac.uk/;

dbSNP, https://www.ncbi.nlm.nih.gov/snp/;

LD-Hub, http://ldsc.broadinstitute.org/; LDlink, https://ldlink.nci.nih.gov/;

PhenoScanner, http://www.phenoscanner.medschl.cam.ac.uk/;

mRnd, https://shiny.cnsgenomics.com/mRnd/.

## Author contributions

Conceptualization: Hao Zhao, Xuening Zhang, Lan Guo, Ciyong Lu

Data curation: Hao Zhao, Lan Guo

Formal analysis: Hao Zhao

Investigation: Xue Han, Wanxin Wang

Methodology: Hao Zhao, Xuening Zhang, Lan Guo

Project administration: Lan Guo, Ciyong Lu

Resources: Xue Han, Yuhua Liao, Huimin Zhang

Software: Hao Zhao, Xuening Zhang

Supervision: Lingjiang Li, Ciyong Lu

Validation: Wenyan Li, Jingman Shi, Wenjian Lai, Lan Guo

Visualization: Xuening Zhang

Writing –original draft: Hao Zhao

Writing – review and editing: Hao Zhao, Xue Han, Lingjiang Li, Xuening Zhang, Yuhua Liao, Huimin Zhang, Wenyan Li, Jingman Shi, Wenjian Lai, Wanxin Wang, Roger S. McIntyre, Kayla M. Teopiz, Lan Guo, and Ciyong Lu

## Acknowledgements

The authors thank the staff and participants of the PGC (Mental Genomics Consortium) for making the GWAS data publicly available.

## Conflict of interest

I have read the journal’s policy and the authors of this manuscript have the following competing interests: Dr. Roger McIntyre has received research grant support from CIHR/GACD/National Natural Science Foundation of China (NSFC); speaker/consultation fees from Lundbeck, Janssen, Alkermes, Mitsubishi Tanabe, Purdue, Pfizer, Otsuka, Takeda, Neurocrine, Sunovion, Bausch Health, Novo Nordisk, Kris, Sanofi, Eisai, Intra-Cellular, NewBridge Pharmaceuticals, Abbvie, Atai Life Sciences. Dr. Roger McIntyre is a CEO of Braxia Scientific Corp. Kayla M. Teopiz has received personal fees from Braxia Scientific Corp. All other authors have no conflicts of interest to disclose.

## Ethical approval

Not required.

## Funding

The author(s) received no specific funding for this work.

## Abbreviations

ADHD: attention-deficit/hyperactivity disorder
ASD: autism spectrum disorder
BIP: bipolar disorder
CI: confidence interval
GWAS: genome-wide association studies
IVW: inverse-variance weighted
LD: linkage disequilibrium
MDD: major depressive disorder
MR: Mendelian randomization
OR: odds ratio
PGC: Psychiatric Genomics Consortium
PTSD: post-traumatic stress disorder
RCTs: randomized clinical trials
SCZ: schizophrenia
SNPs: single nucleotide polymorphisms

## Supporting information

**S1 STROBE-MR-Checklist**.

(DOCX)

**S1 Table. The summary information of the studies used for genetic instrumental variables extraction of absolute circulating antioxidants**.

(DOCX)

**S2 Table. The summary statistics of the six major psychiatric disorders**.

(DOCX)

**S3 Table. The statistical power in Mendelian randomization study on association of diet-derived circulating antioxidants with risk of six major psychiatric disorders**.

(DOCX)

**S4 Table. The causal effect estimates of the associations between genetic instrumental variables for absolute blood antioxidants and risk of six major psychiatric disorders**.

(DOCX)

**S5 Table. The causal effect estimates of the associations between genetic instrumental variables for antioxidant metabolite concentrations and risk of six major psychiatric disorders**.

(DOCX)

**S6 Table. The sensitivity MR analysis results of the causal effects of absolute α-tocopherol levels on SCZ from other GWAS studies**.

(DOCX)

**S1 Figure. Scatter plots of the SNP effect of absolute α-tocopherol levels and SCZ in the primary MR analysis**.

(DOCX)

**S2 Figure. Forest plot of primary MR analysis of absolute α-tocopherol levels on SCZ**.

(DOCX)

**S3 Figure. Plot of primary MR leave-one-out analysis of absolute α-tocopherol levels on SCZ**.

(DOCX)

## Notes

### Author Declarations

Because all analyses here in are based on publicly available summary data, no ethical approval from an institutional review boardwas required for this study.

